# Prolactin-increasing antipsychotics use and reduced risk of prostate cancer: a population-wide cohort and nested case-control study

**DOI:** 10.1101/2025.10.28.25339029

**Authors:** Qi Sun, Yifang Huang, Yuqi Hu, Wenxin Tian, Song Song, Rachel Yui Ki Chu, Xinya Mu, Min Fan, Franco WT Cheng, Wenlong Liu, Cuiling Wei, Lingyue Zhou, Boyan Liu, Zijie Xu, Esther Wai Yin Chan, Ian Chi Kei Wong, Francisco Tsz Tsun Lai

**Author notes:** Correspondence Dr. Francisco Tsz Tsun Lai, PhD, L2-26A, Laboratory Block, Li Ka Shing Faculty of Medicine, The University of Hong Kong, 21 Sassoon Road, Pokfulam, Hong Kong SAR.

## Abstract

**Objective:** This study aims to investigate whether users of prolactin-increasing antipsychotics exhibit a lower risk of prostate cancer compared to those using prolactin-sparing antipsychotics.

**Methods:** Using territory-wide electronic health records from Hong Kong, we conducted a retrospective cohort study along with a nested case-control analyses involving adults who were prescribed antipsychotics and had no history of cancer, organ transplantation, or sexually transmitted infections. Baseline characteristics included demographics, comorbidities, and co-medications. The outcome of interest was incident prostate cancer. Poisson regression, incorporating inverse probability of treatment weighting (IPTW) and multivariable adjustment, were used to estimate incidence rate ratios (IRR) in the cohort analyses. A conditional logistic regression model was applied in the nested case-control analyses to calculate odds ratios (OR) associated with longer-term use of prolactin-increasing antipsychotics, compared to short-term use or non-use.

**Results:** The cohort study included 81,177 individuals in the primary analyses. Users of prolactin-increasing antipsychotics showed a halved risk of prostate cancer compared to the prolactin-sparing group, with an adjusted IRR (aIRR) of 0.410 (95% CI: 0.237–0.710). In the nested case-control analyses, 291 cases were identified. A similarly reduced risk was observed among those who had used prolactin-increasing antipsychotics for 1–5 years, with an adjusted OR (aOR) of 0.569 (95% CI: 0.356–0.909). This association is more pronounced in people under the age of 75 but not among those aged above.

**Conclusion:** Prolactin-increasing antipsychotics were associated with a markedly lower risk of prostate cancer. If the findings are further substantiated, this potential benefit from prolactin-increasing antipsychotics should be considered.

## INTRODUCTION

Prostate cancer is ranked the fourth most frequently diagnosed cancer globally in 2022, accounting for 7.3% of all cancers across 118 out of 185 countries^1^, and represents the second leading cause of cancer-related mortality among men in the United States.^2^ Over the past decade, the incidence of prostate cancer in the U.S. has risen by 3.0% annually, with the highest proportion of new cases occurring among men aged 65 to 74 years.

Among other risk factors, previous research has estimated that men with low free testosterone levels exhibit as much as one-fourth lower risk of prostate cancer ^3^. Testosterone reduction is thus an established strategy for prostate cancer prevention and treatment, typically by the use of 5-alpha-reductase inhibitors.^4^ In fact, lower free testosterone levels can also be associated with elevated prolactin levels,^5^ suggesting a potential avenue for novel drug repurposing in prostate cancer prevention.

Prolactin-increasing antipsychotics, typically used for people with severe mental illness, act primarily through dopamine receptor antagonism in the pituitary gland, thereby inhibiting dopamine-mediated suppression of prolactin secretion.^6^ Prolonged use of these agents can result in a continued heightened level of prolactin, which may subsequently lead to continuously suppressed testosterone levels.^5^ Preclinical studies have shown a potential impact of haloperidol, a prolactin-increasing antipsychotic drug, on reducing prostate cancer risks.^7^ Nevertheless, no direct comparative study has assessed the risk of prostate cancer between users of prolactin-increasing and prolactin-sparing antipsychotics. The potential protective effect of prolactin-increasing antipsychotics remains unexplored in real-world settings, and its clinical implications unclear.

In this study, we leveraged a territory-wide longitudinal public healthcare database to conduct a retrospective cohort study along with nested case-control analyses to evaluate the association between prolactin-increasing antipsychotic use and the risk of prostate cancer. Our primary objective was to determine whether users of prolactin-increasing antipsychotics experience a lower risk of prostate cancer compared to those prescribed prolactin-sparing antipsychotics (retrospective cohort study). Additionally, we aimed to examine whether longer duration of prolactin-increasing antipsychotic use is associated with a further reduction in prostate cancer risk (nested case-control study).

## METHODS

### Data source

This retrospective cohort study with a nested case-control analyses utilized territory-wide electronic health records from the Hong Kong Hospital Authority (HA), covering the period from 1 January 2004 to 31 December 2023. The HA is the primary provider of public healthcare services in Hong Kong, managing all public hospitals and most public outpatient clinics, and serving a population of over 7.5 million residents.

The dataset included anonymized demographic details, prescription records, mortality information, and clinical diagnoses. Diagnostic records were available from 1 January 1993 onward to capture sufficient medical history, while other data were collected from 1 January 2004 onward. All diagnoses were recorded by registered clinicians using the International Classification of Diseases, Ninth Revision, Clinical Modification (ICD-9-CM), with precise timestamps. Death records, including causes of death coded in ICD-10, were obtained from the official city-wide death registry. All records were linkable via a unique anonymous patient identifier assigned to each resident.

### Study design and participants

To estimate the average treatment effect (ATE) on prostate cancer risk comparing users of prolactin-increasing versus prolactin-sparing antipsychotics, we conducted a retrospective cohort study. Cumulative exposure patterns were additionally examined in a nested case– control analysis.

### Retrospective cohort study

This retrospective cohort included male patients aged 18 years or older in Hong Kong who initiated treatment with either prolactin-increasing or prolactin-sparing antipsychotics^8-10^ between 1 January 2004 and 31 December 2023. Patients were classified into exposure groups according to the class of antipsychotic initiated, and the date of the first qualifying antipsychotic prescription was designated as the index date (see eFigure 1). To implement a new-user design, we excluded individuals with any recorded use of antipsychotics before 1 January 2006. Additional exclusions were a history of cancer (other than non-melanoma skin cancer, to ensure incident case ascertainment), organ transplantation, or sexually transmitted diseases (STDs) before the index date. Enrolment window is till 31 December 2018, so all included participants were eligible for at least five years of follow-up after the index date.

Patients were followed from the index date until the occurrence of prostate cancer diagnosis, death from other causes (except prostate cancer caused death), discontinuation of the initial antipsychotic type (defined as an interval of non-use exceeding 90 days or switching antipsychotics group), or the end of the study period (31 December 2023), whichever occurred first.

The primary outcome was the first diagnosis of prostate cancer occurring at least one year after the index date. A one-year lag after cohort entry was applied to reduce reverse causation, given the limited biological plausibility that very recent exposure would influence cancer development. In addition, a one-year lag after discontinuation was implemented to reflect a plausible latency window for drug-related cancer initiation or promotion.^11,12^

The primary exposure was prolactin-increasing antipsychotics being compared to prolactin-sparing antipsychotics (see eTable 3).^10,13,14^ Demographic characteristics, comorbidities, and prior medication use related to prostate cancer risk were considered as covariates. Comorbidities included cardiovascular diseases, metabolic syndrome, non-alcoholic fatty liver disease, asthma, chronic obstructive pulmonary disease, diabetes, alcohol misuse, tobacco misuse, periodontitis, prostatitis, ulcerative colitis, infertility, Sjögren’s syndrome, dementia, depressive disorder, schizophrenia, bipolar disorder, anxiety disorder, suicide attempt, and vasectomy (see eTable 5 with corresponding ICD-9-CM or ICD-10 codes). Prior medication classes included acid-suppressive drugs, aspirin, selective serotonin reuptake inhibitors (SSRIs), angiotensin receptor blockers, angiotensin-converting enzyme inhibitors, calcium channel blockers, non-steroidal anti-inflammatory drugs, diuretics, lipid-regulating drugs, beta-blockers, insulin, metformin, thiazolidinediones, and allopurinol (see eTable 4). Cumulative duration of these drugs was categorized as <1 year, 1–5 years, and ≥5 years. In the cohort analyses, covariates were ascertained prior to the index date and considered in both primary and sensitivity analyses.

Poisson regression with robust variance and an offset for follow-up time was used to estimate incidence rate ratios (IRRs) with 95% confidence intervals (CIs). To mitigate potential confounding from the covariates, propensity scores were estimated via logistic regression and used to construct inverse probability of treatment weights (IPTW). Overlap IPTW was applied to enhance comparability between exposure groups. Covariate balance was assessed using standardized mean differences (SMDs), with SMD < 0.01 indicating adequate balance. As an alternative statistical estimation approach, Cox regression was conducted to estimate the hazard ratios (HRs) of prostate cancer with 95% CIs. To address potential bias from competing risks, we fitted cause-specific Cox proportional hazards models to estimate cause-specific hazard ratios (HRs). Subgroup analysis by age (below versus 75+) was also conducted to observe age differences of the associations. Both weighted and unweighted (crude) estimates were generated in all the cohort analyses.

### Nested case-control study

We conducted a nested case–control analysis on the retrospective cohort to evaluate the association of cumulative exposure to prolactin-increasing antipsychotics with prostate cancer risk. Cases were men with an initial diagnosis of prostate cancer between January 2006 and December 2023; the diagnosis date served as the index date (see eFigure 4). Eligible cases were aged 18–85 years at diagnosis and had at least five years of medication records prior to the index date. Individuals with a prior cancer diagnosis (except non-melanoma skin cancer), a history of organ transplant, or any history of sexually transmitted diseases (STD) were excluded. A total of 291 cases (9.2%) were identified from an eligible pool of 3,150 men.

For each case of prostate cancer patient, up to ten age-matching (±1 year) controls (without prostate cancer at the time the first prostate cancer diagnosis was diagnosed in the case patient) were selected using risk-set sampling.^15^. The index date for the controls was mapped from the case patients’ duration of prostate cancer-free periods since their first antipsychotic prescription. In other words, at the index date, cases and controls had been observed in the cohort for the same amount of time. The same exclusion criteria were applied to controls, who were allowed to subsequently become cases (>1 year later). Each control could be selected no more than four times for different cases.

Antipsychotic exposure was ascertained from 2004 up to one year before each individual’s index date to allow for a biologically relevant latency period.^11,12^ Exposure was defined as the cumulative duration of antipsychotic use (in days) and categorized as ≤1 year, 1–5 years, and ≥5 years. Periods of prolactin-sparing antipsychotic use were restricted to times when these agents were prescribed without concomitant prolactin-increasing antipsychotics. Conversely, any combination regimen that included a prolactin-increasing antipsychotic—whether or not a prolactin-sparing agent was co-prescribed—was classified as prolactin-increasing exposure. Because aripiprazole, a partial dopamine receptor agonist with additional serotonergic activity, may attenuate prolactin elevation when co-administered with prolactin-increasing antipsychotics,^16-18^ primary analyses further subdivided exposure into four mutually adjusted categories: i. prolactin-increasing antipsychotic use; ii. prolactin-sparing antipsychotic use; iii. prolactin-increasing antipsychotic use with concomitant aripiprazole; and iv. prolactin-sparing antipsychotic use with concomitant aripiprazole. Each category was included simultaneously in the final regression model.

Covariates and their measurement are the same as described for the cohort study. Covariates were ascertained up to one year before the index date and incorporated in both main and sensitivity analyses. Conditional multivariable logistic regression was used to estimate odds ratios (ORs) and 95% confidence intervals (CIs), with ≤1 year of use (including never-users in either prolactin-increasing or prolactin-sparing group) as the reference category. Subgroup analyses stratified by age (<75 and ≥75 years) were conducted as well, consistent with the cohort study.

Analyses were conducted in R (version 4.4.0). Participants with missing demographic information were excluded using complete-case analysis. Statistical significance was evaluated with two-sided tests at α = 0.05. Results were independently cross-checked by QS and YH to ensure analytical accuracy.

## RESULTS

A total of 443,176 patients were prescribed antipsychotics (as classified under B.N.F. sections 4.2.1 and 4.2.2, excluding lithium) between 1 January 2004 and 31 December 2023. After excluding females, individuals under 18 years of age at first antipsychotic use, those with less than five years of follow-up, patients who used both prolactin-increasing and prolactin-sparing antipsychotics concomitantly or initiated treatment with aripiprazole, as well as those with a history of cancer, sexually transmitted diseases, or organ transplantation, a total of 81,177 patients were included in the cohort study (Figure 1). Baseline characteristics are summarized in Table 1.

**Figure 1.**
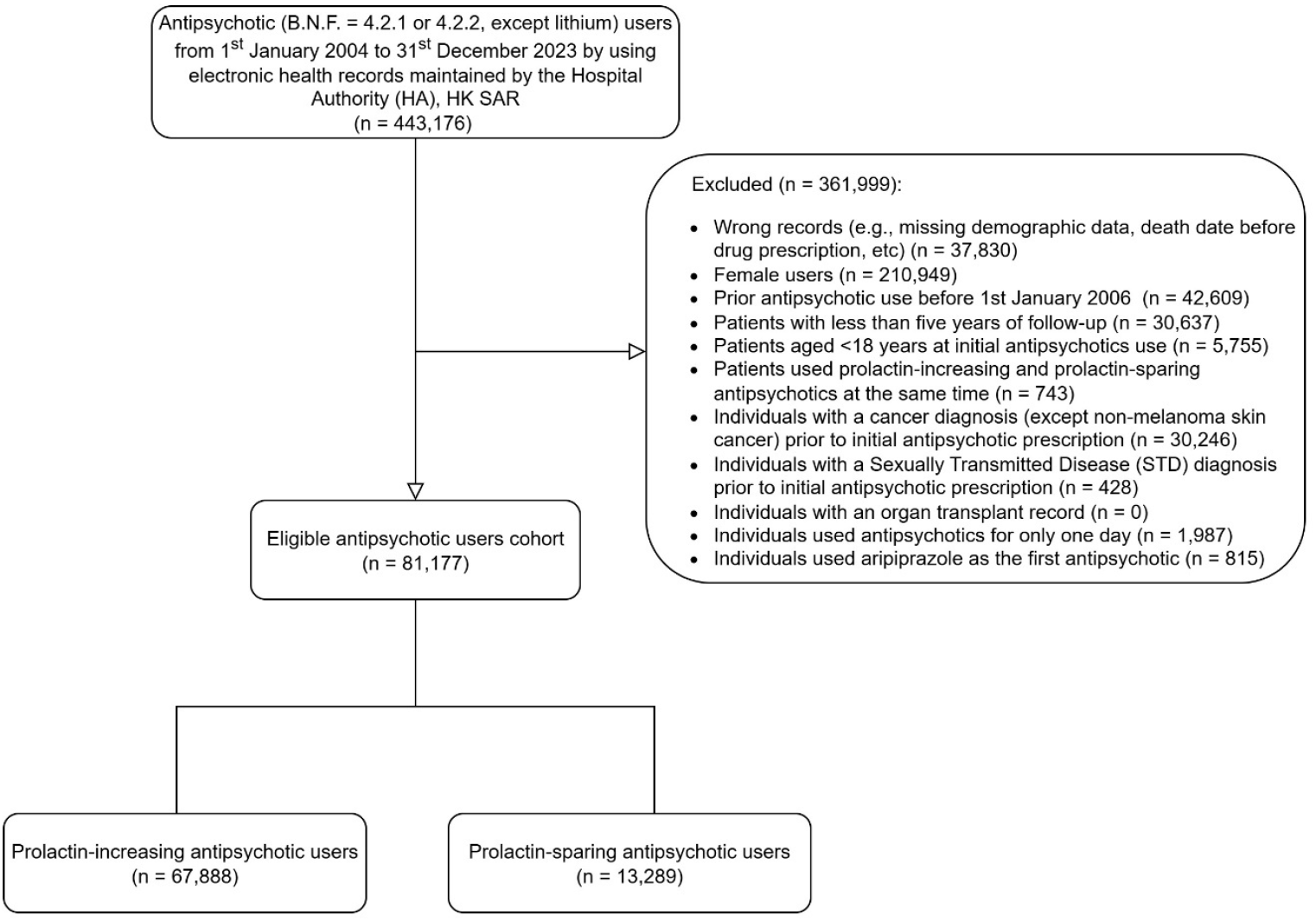
Flowchart of cohort analyses

**Table 1.**
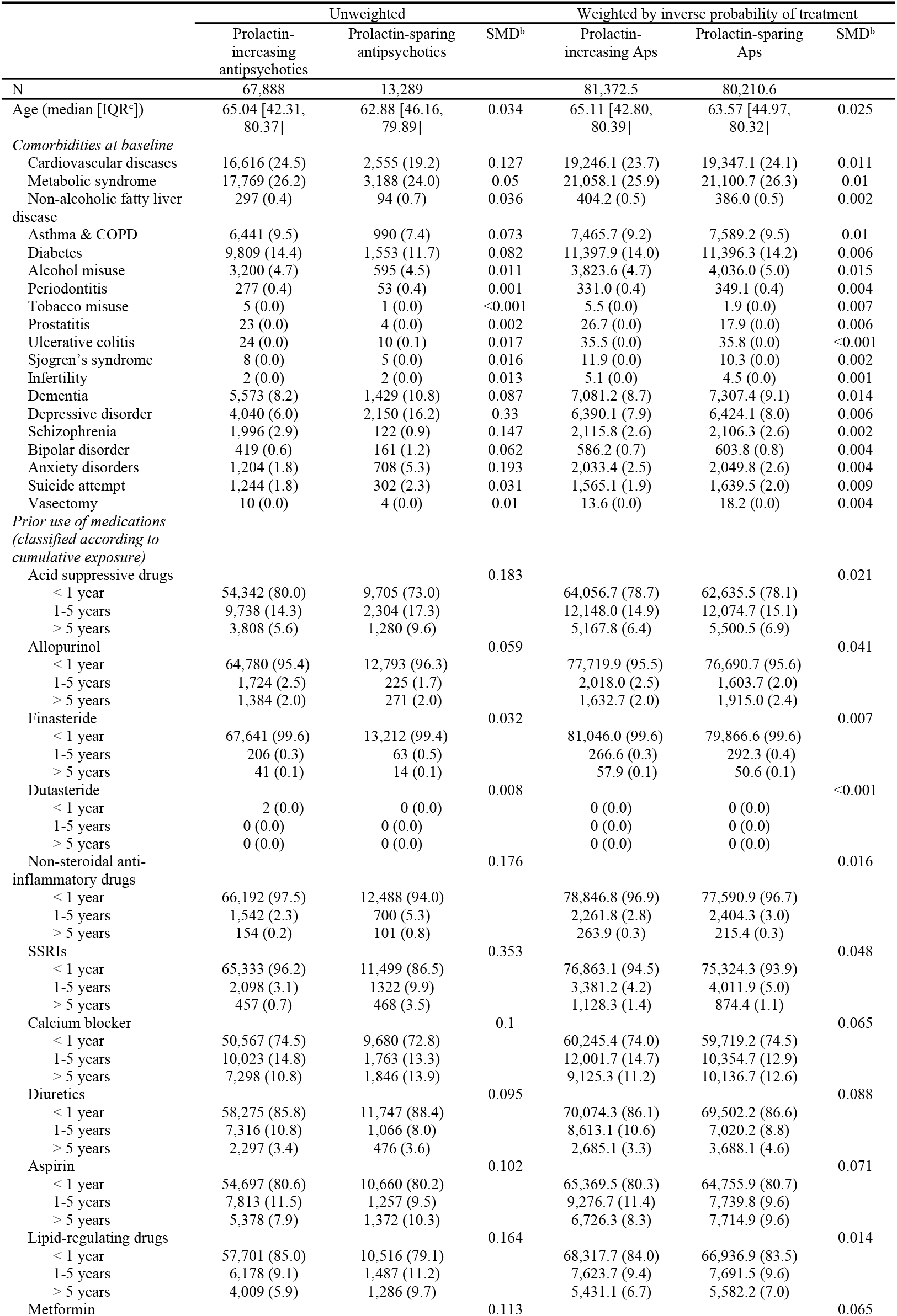
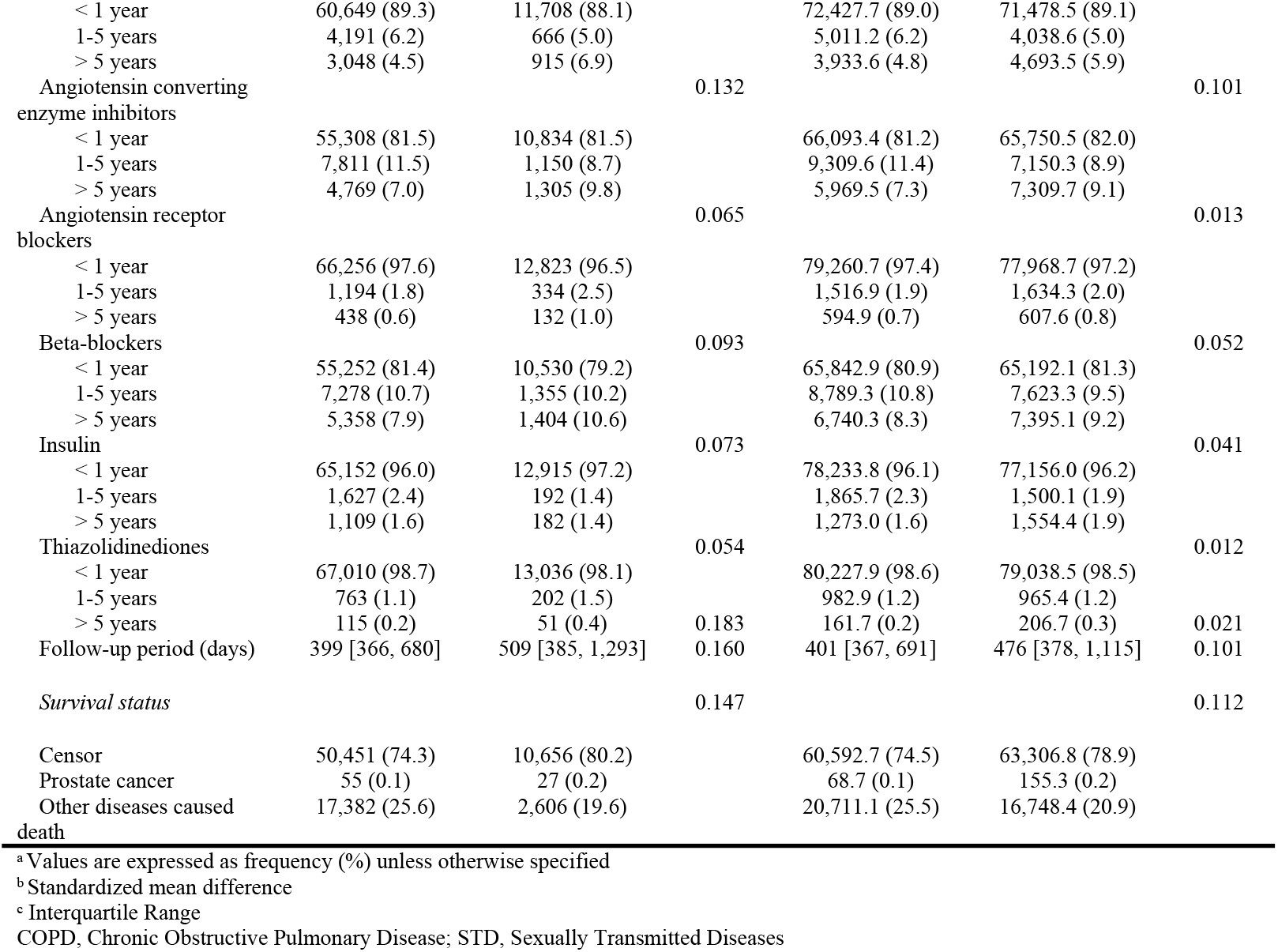
Baseline characteristics in cohort analyses^a^.

Among the 81,177 patients, 67,888 (median age: 65.04 years, Interquartile Range (IQR): 42.31-80.37 years) were users of prolactin-increasing antipsychotics, with a median follow-up of 399 days (IQR: 366-680 days), and 13,289 (median age: 62.88 years, IQR: 46.16-79.89 years) used prolactin-sparing antipsychotics, with a median follow-up of 509 days (IQR: 385-1,293 days). During the follow-up, 55 and 27 incident cases of prostate cancer were identified in the prolactin-increasing and prolactin-sparing groups, respectively.

Figure 2 presents the cumulative hazard of prostate cancer overtime for both groups. During the first one to five years of follow-up, the prolactin-increasing group exhibited a significantly lower cumulative hazard of prostate cancer, with a 46.8% risk reduction compared to the prolactin-sparing group. The adjusted incidence rate ratio (aIRR) was 0.410 (95% CI: 0.237– 0.710), and the adjusted hazard ratio (aHR) was 0.532 (95% CI: 0.301–0.942) in Table 2.

**Table 2.** Association between prolactin-increasing antipsychotics and the risk of prostate cancer with 1-year lag window.

|  | Cohort size | Average Follow-up days | Number of prostate cancers | Crude IRR with 95% CI | Doubly robust adjusted* IRR with 95% CI | Doubly robust adjusted* IRR with 95% CI in 1-5 years follow-up period | Doubly robust adjusted* HR with 95% CI | Doubly robust adjusted* HR with 95% CI in 1-5 years follow-up period | Doubly robust adjusted* cause-specific hazard rate | Doubly robust adjusted* cause-specific hazard rate in 1-5 years follow-up period |
| --- | --- | --- | --- | --- | --- | --- | --- | --- | --- | --- |
| Overall | 81,177 | 847 | 83 |  |  |  |  |  |  |  |
| Prolactin-increasing Antipsychotic | 67,888 | 819 | 55 | 0.482 (0.304, 0.764) | 0.646 (0.397, 1.051) | 0.410 (0.237, 0.710) | 0.793 (0.471, 1.334) | 0.532 (0.301, 0.942) | 0.793 (0.471, 1.334) | 0.532 (0.301, 0.942) |
| Prolactin-sparing Antipsychotic | 13,289 | 990 | 27 | Ref. | Ref. | Ref. | Ref. | Ref. | Ref. | Ref. |

\*: Doubly robust adjusted: after using inverse probability treatment weight (IPTW) to balance the baseline characteristics distribution between prolactin-increasing antipsychotics and prolactin-sparing antipsychotics group, we adjusted these confounders in the final model with IPTW as well.

**Figure 2.**
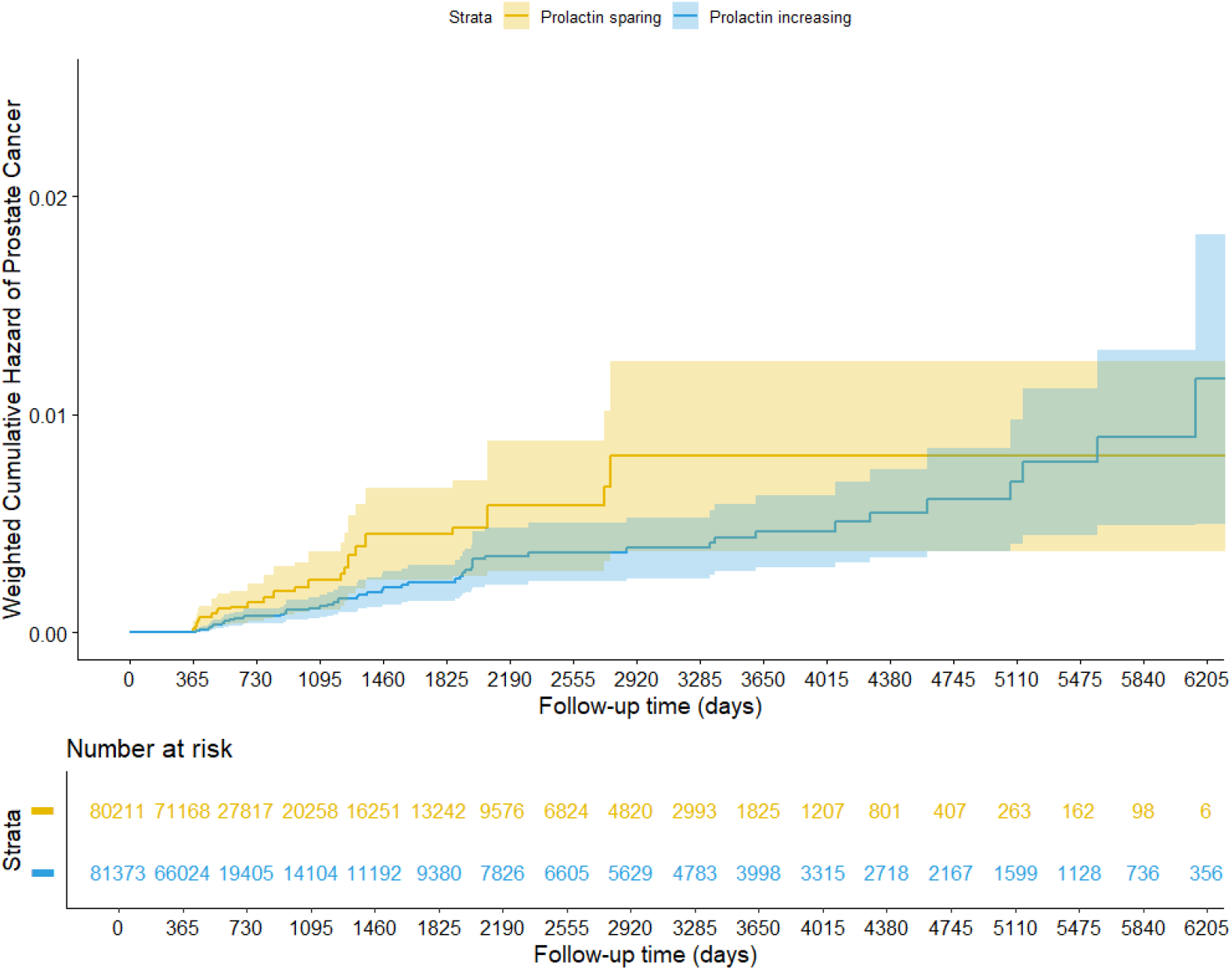
Weighted cumulative hazard plot of prostate cancer.

This protective association was more pronounced in patients younger than 75 years, with an adjusted IRR of 0.436 (95% CI: 0.227–0.837). In contrast, no significant association was observed among patients aged 75 years or older (IRR: 1.160, 95% CI: 0.526–2.560; see eTable 1, eFigure 2, and eFigure 3).

In the nested case-control analyses, 291 incident cases of prostate cancer were identified and matched to 2,859 controls from the eligible cohort (Figure 3). The median age was approximately 75 years in both groups (cases: median 75.01 years, IQR 68.30–80.40; controls: median 74.89 years, IQR 68.13–80.33). The distributions of prolactin-increasing antipsychotics, prolactin-sparing antipsychotics, and concomitant use of these classes with aripiprazole were comparable between cases and controls (Table 3). Specifically, prolactin-sparing antipsychotics were used for more than five years by 3.4% of cases and 1.7% of controls, while prolonged use (≥5 years) of prolactin-increasing antipsychotics was observed in 5.5% of cases and 7.2% of controls.

**Table 3.**
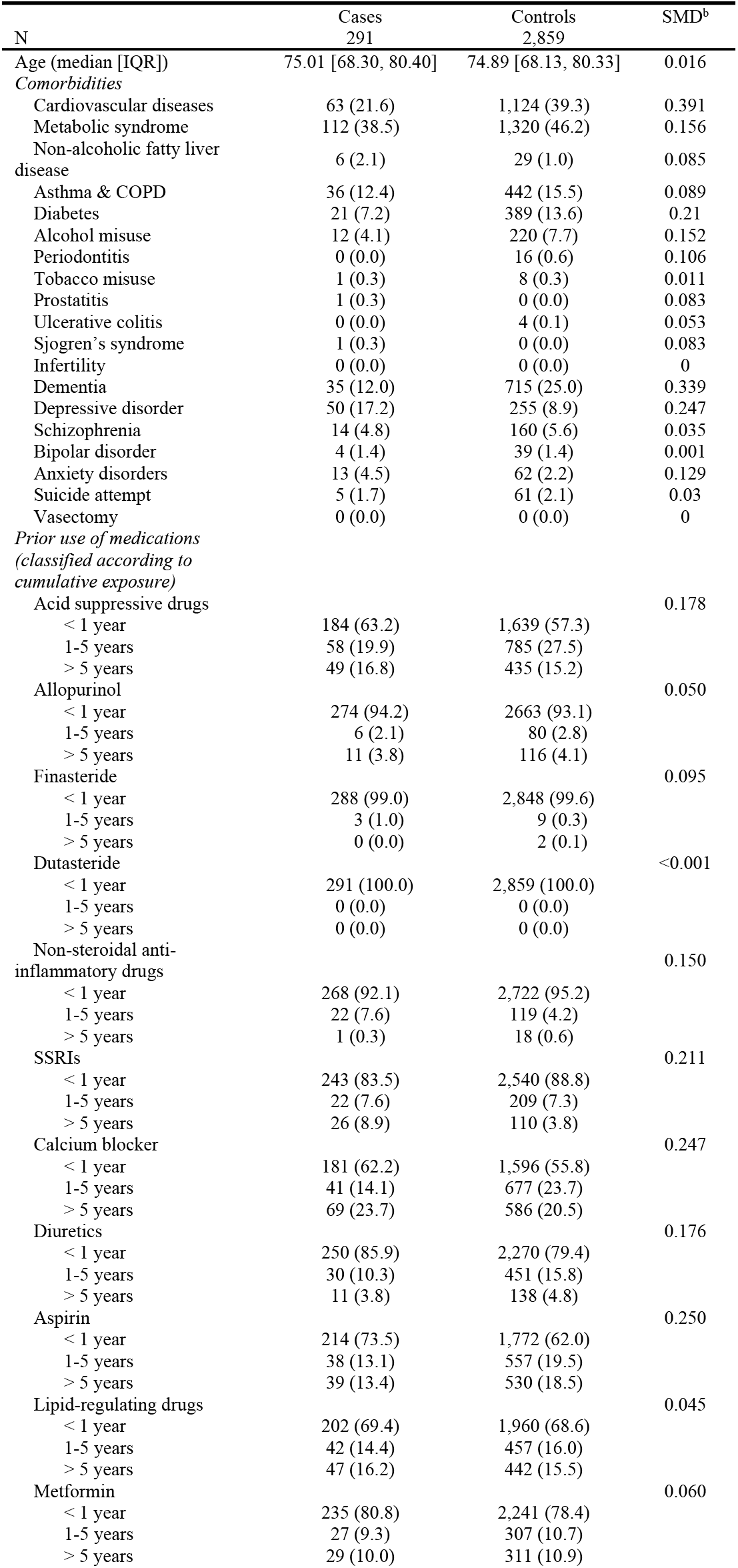
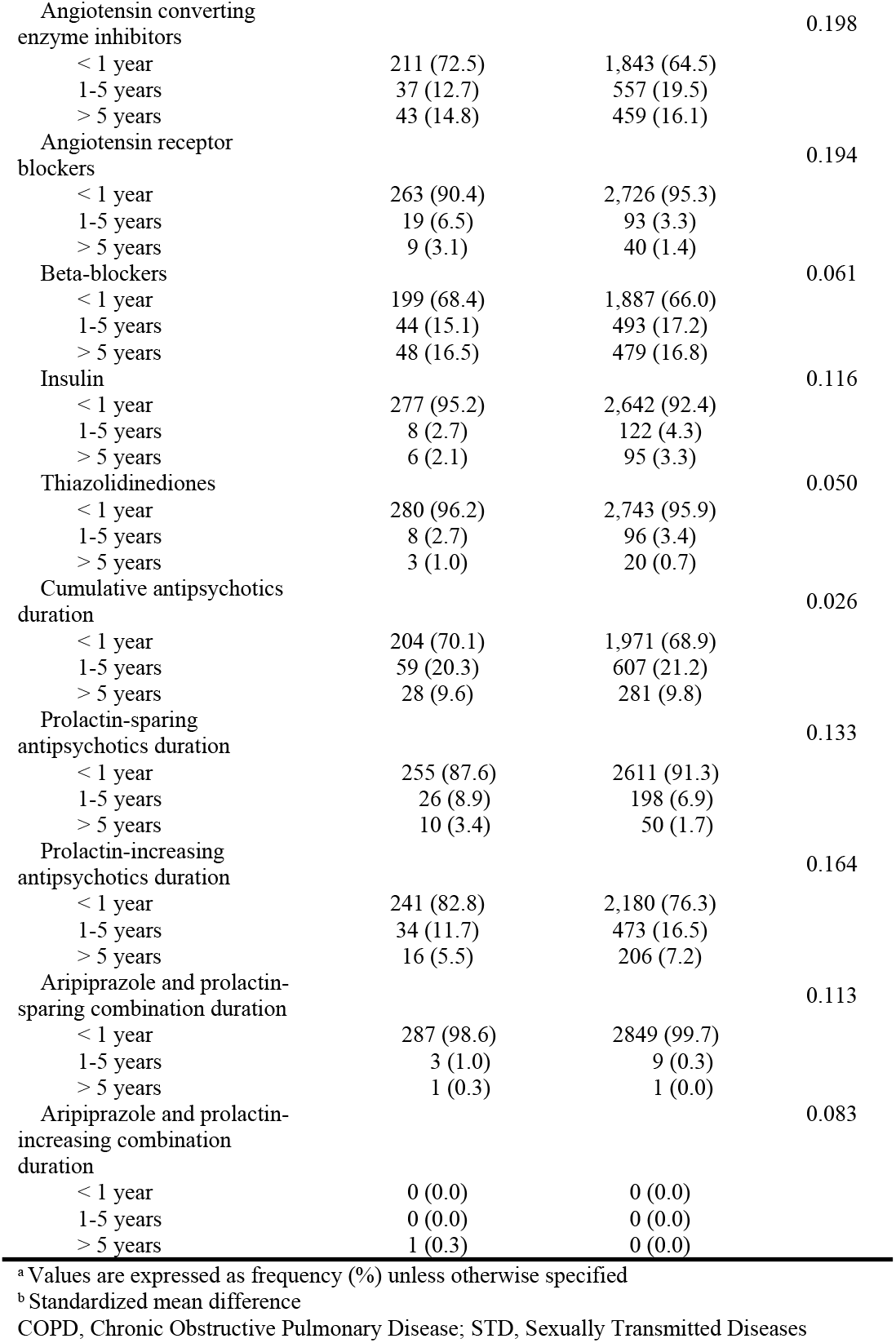
Characteristics of men with prostate cancer (cases) and without prostate cancers (controls)

**Figure 3.**
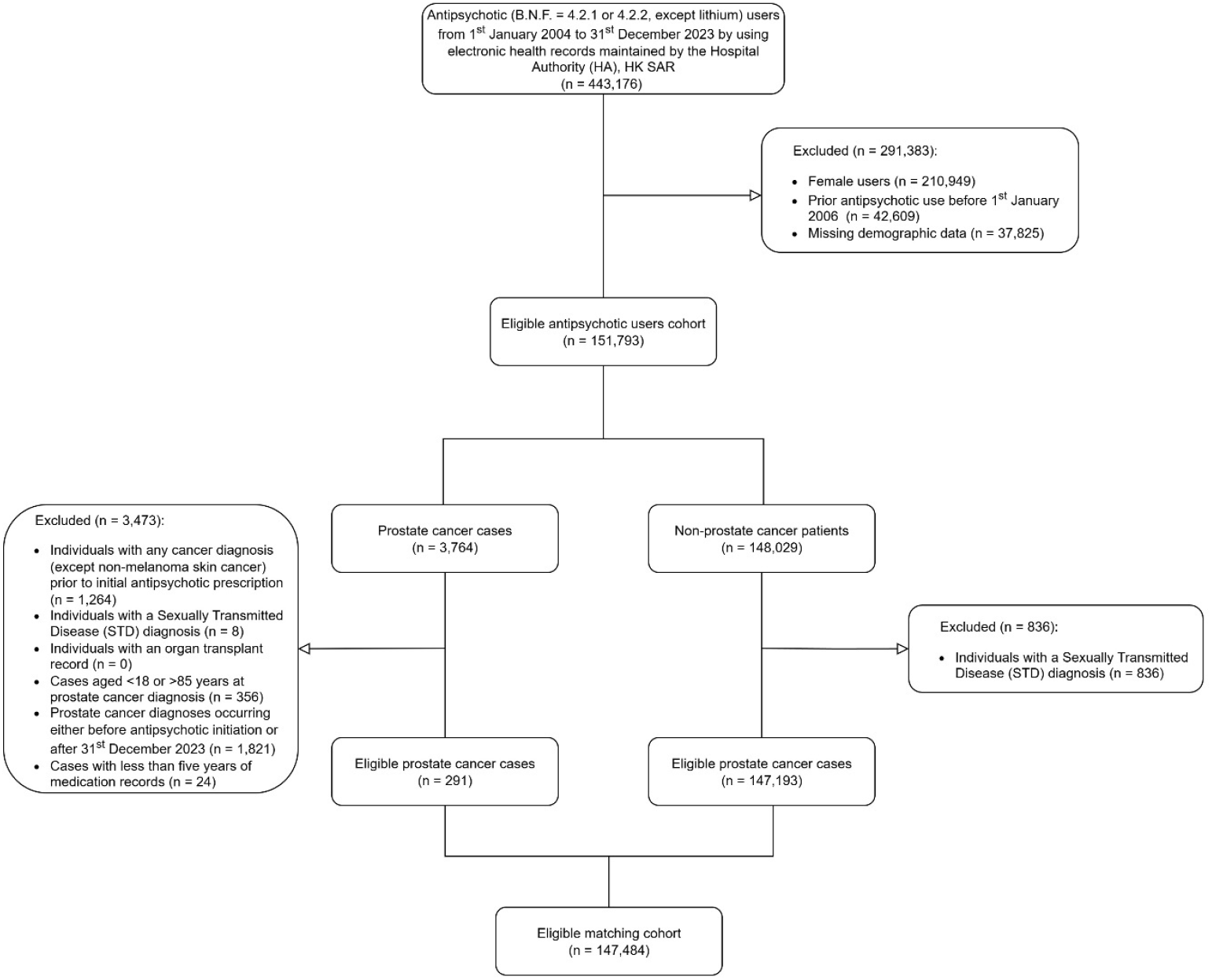
Flowchart of nested case-control analyses

In Table 4, compared to minimal exposure (reference category), use of prolactin-increasing antipsychotics for 1–5 years was associated with a significantly lower risk of prostate cancer, with an adjusted odds ratio (aOR) of 0.569 (95% CI: 0.356–0.909; p = 0.014). In contrast, use beyond five years did not show a statistically significant association (aOR: 0.575; 95% CI: 0.288–1.147). For prolactin-sparing antipsychotics, neither medium-term (1–5 years; aOR: 0.905; 95% CI: 0.537–1.527) nor long-term use (≥5 years; aOR: 1.428; 95% CI: 0.633–3.222) was significantly associated with prostate cancer risk.

**Table 4.** Association between cumulative prolactin-increasing antipsychotics exposure duration and the risk of prostate cancer with 1-year lag window.

|  | Case<br>N = 291 | Control<br>N = 2,859 | Unadjusted OR<br>95% CI | Adjusted* OR with 95%<br>CI |
| --- | --- | --- | --- | --- |
| Duration of any antipsychotics use |  |  |  |  |
| < 1 year | 204 (70.1) | 1,971 (68.9) | Ref. | Ref. |
| 1-5 years | 59 (20.3) | 607 (21.2) | 0.885 (0.608, 1.287) | 0.798 (0.525, 1.213) |
| > 5 years | 28 (9.6) | 281 (9.8) | 0.842 (0.493, 1.439) | 0.794 (0.436, 1.445) |
| Duration of prolactin-increasing antipsychotics use† |  |  |  |  |
| < 1 year | 241 (82.8) | 2,180 (76.3) | Ref. | Ref. |
| 1-5 years | 34 (11.7) | 473 (16.5) | 0.562 (0.370, 0.854) | 0.569 (0.356, 0.909) |
| > 5 years | 16 (5.5) | 206 (7.2) | 0.621 (0.329, 1.172) | 0.575 (0.288, 1.147) |
| Duration of prolactin-sparing antipsychotics use‡ |  |  |  |  |
| < 1 year | 255 (87.6) | 2611 (91.3) | Ref. | Ref. |
| 1-5 years | 26 (8.9) | 198 (6.9) | 1.228 (0.771, 1.955) | 0.905 (0.537, 1.527) |
| > 5 years | 10 (3.4) | 50 (1.7) | 1.909 (0.892, 4.087) | 1.428 (0.633, 3.222) |

\*: Adjusted for: age at prostate cancer diagnose dates in cases or matching date of controls, comorbidities, and prior use of medications before 1 year prior to index date. Furthermore, prolactin-increasing, prolactin-sparing antipsychotics use, aripiprazole and prolactin-increasing combination use, and aripiprazole and prolactin-sparing combination use are adjusted for each other. † when excluding time with concomitant use of aripiprazole. ‡ when excluding time with concomitant use of aripiprazole or prolactin-increasing antipsychotics.

Sensitivity analyses stratified by age revealed a consistent protective relationship associated with 1–5 years of prolactin-increasing antipsychotic exposure. In patients younger than 75 years, the aOR was 0.507 (95% CI: 0.268–0.961), corresponding to a 49.3% risk reduction, while in those aged 75 years or older, the aOR was 0.489 (95% CI: 0.227–1.050), which did not reach statistical significance.

## DISCUSSION

To the best of our knowledge, this is the first territory-wide retrospective cohort and nested case-control study to evaluate the association between prolactin-increasing antipsychotic use and the risk of prostate cancer, using large-scale electronic health records and an active-comparator design (prolactin-sparing antipsychotic users). Robust and consistent findings from both analyses suggest a potential clinically significant protective association of prolactin-increasing antipsychotics against prostate cancer.

Apart from the obvious absence of randomization, two key limitations of this study should be acknowledged. Firstly, Our dataset did not include information on cancer stage or histological type of prostate cancer. Besides, details on precise dosage and frequency of antipsychotic use were not available, which could have offered a more refined measure of exposure than duration alone, particularly in the nested case-control analysis. Future studies incorporating prescription records with dosage information—as typically recorded by clinicians and psychiatrists following treatment guidelines—would strengthen exposure assessment. Second, to enhance the generalizability of our findings, further research in other populations is warranted, given the known variations in prostate cancer risk across ethnicities, especially in African American.^19^

In the context of the literature, our findings complement those from a U.S. case-control study that investigated the anticancer potential of antipsychotics based on sigma-1 receptor affinity,^20^ which reported an aOR of 0.38 (95% CI: 0.14–1.01) for haloperidol (a sigma-1 antagonist, an antipsychotic with high prolactin-inducing propensities) use beyond one year, and an aOR of 0.80 (95% CI: 0.66–0.98) for risperidone, quetiapine, and olanzapine (non-sigma-1 agents). Mechanistic research has suggested that haloperidol’s dual affinity for dopamine D2 and sigma-1 receptors may suppress androgen receptor (AR) and AR splice variant (ARV) activity in prostate cancer models.^21^ However, no real-world studies so far have employed an active-comparator design to compare a wider range of prolactin-increasing drugs versus prolactin-sparing drugs and address potential indication bias as well as multivariable adjustments for key confounders such as underlying mental illnesses.

The potential protective effect of prolactin-increasing antipsychotics may be attributed to hyperprolactinemia-induced reduction in free testosterone levels. This aligns with evidence indicating that lower circulating testosterone is associated with a decreased risk of prostate cancer.^22^ For instance, Eleanor et al. reported an aOR of 0.77 (95% CI: 0.69–0.86) for men in the lowest decile of free testosterone compared to those in higher deciles.^3,23^ The absence of a significant effect in older patients (≥75 years) may reflect the stronger influence of age and ethnic factors on prostate cancer risk in this population.

Current pharmacological strategies for prostate cancer prevention primarily involve 5-alpha-reductase inhibitors such as finasteride and dutasteride, which reduce dihydrotestosterone (DHT) levels and thereby mitigate cancer risk.^4^ If our findings are further substantiated, there may be implications for guidelines concerning the management of people with severe mental illness at an elevated risk of prostate cancer. Clinicians could consider prolactin-increasing antipsychotics for carefully selected patients to reduce the risk of prostate cancer along with the intended antipsychotic treatment goal. Such recommendations would need to also be jointly considered with the other potential side effects of prolactin-increasing drugs, such as bone fractures,^24^ cardiovascular events,^25^ neuropathy^26^, psoriasis,^27^ weight gain,^28^ and cognitive decline.^29^ These risks are particularly relevant among older men.

## CONCLUSION

This study demonstrates that the use of prolactin-increasing antipsychotics is associated with a reduced risk of prostate cancer compared with prolactin-sparing antipsychotics in men, particularly among those under 75 years of age. Further research is needed to substantiate these findings and to explore the underlying mechanisms to guide potential clinical recommendations that take advantage of this potential benefit.

## Data Availability

Data will not be accessible to others as the data custodian has not granted permission.

## Contributors’ Statement

Qi Sun and Francisco Lai contributed the conception of this work. The study was designed by all authors. Specifical thanks to Dr. Min Fan and Dr. Franco W.T. CHENG in study design consultant. The study design, baseline characteristics, and codes were doubly checked by YF Huang. Qi Sun contributed to the acquisition and analyses of the data, and all authors interpreted the data. Qi Sun and Francisco Lai drafted the manuscript. Every contributor critically reviewed the manuscript for essential intellectual content, endorsed the final version for publication, and accepted responsibility for all components of the work. Qi Sun is the first author and Francisco Lai is the corresponding author. Francisco Lai had full access to all the data in this work, and all authors took ultimate responsibility for the decision to submit this manuscript for publication.

## ACKNOWLEDGEMENTS

This research received no external funding.

## ETHICS APPROVAL

The Institutional Review Board of the University of Hong Kong/Hospital Authority Hong Kong West Cluster (HKU/HA HKW IRB, reference number: UW 20-113) approved this study. Informed consent was not required, as the database was anonymized, and the data collection process complied with data privacy regulations.

## Data Availability

Data will not be accessible to others as the data custodian has not granted permission.

## References

1. Bray F, Laversanne M, Sung H, et al. Global cancer statistics 2022: GLOBOCAN estimates of incidence and mortality worldwide for 36 cancers in 185 countries. CA Cancer J Clin. 2024;74(3):229–263.

2. Siegel DA, O’Neil ME, Richards TB, Dowling NF, Weir HK. Prostate Cancer Incidence and Survival, by Stage and Race/Ethnicity - United States, 2001-2017. MMWR Morb Mortal Wkly Rep. 2020;69(41):1473–1480.

3. Watts EL, Appleby PN, Perez-Cornago A, et al. Low Free Testosterone and Prostate Cancer Risk: A Collaborative Analysis of 20 Prospective Studies. European Urology. 2018;74(5):585–594.

4. Chau CH, Figg WD. Revisiting 5α-reductase inhibitors and the risk of prostate cancer. Nat Rev Urol. 2018;15(7):400–401.

5. Redman B, Kitchen C, Johnson KW, Bezwada P, Kelly DL. Levels of prolactin and testosterone and associated sexual dysfunction and breast abnormalities in men with schizophrenia treated with antipsychotic medications. J Psychiatr Res. 2021;143:50–53.

6. González-Rodríguez A, Labad J, Seeman MV. Antipsychotic-induced Hyperprolactinemia in aging populations: Prevalence, implications, prevention and management. Progress in Neuro-Psychopharmacology and Biological Psychiatry. 2020;101:109941.

7. Friedman GD, Habel LA, Achacoso N, et al. Haloperidol and Prostate Cancer Prevention: More Epidemiologic Research Needed. Perm J. 2020;24.

8. Lai FTT, Tian W, Hu Y, et al. Incidence of gynecological cancers following prolactin-increasing antipsychotic use: a population cohort study. World Psychiatry. 2025;24(3):447–448.

9. Hope JD, Keks NA, Copolov DL. Association between long-term use of prolactin-elevating antipsychotics in women and the risk of breast cancer: What are the clinical implications? Australas Psychiatry. 2023;31(2):205–208.

10. Peuskens J, Pani L, Detraux J, De Hert M. The effects of novel and newly approved antipsychotics on serum prolactin levels: a comprehensive review. CNS Drugs. 2014;28(5):421–453.

11. Pottegård A, Friis S, Stürmer T, Hallas J, Bahmanyar S. Considerations for Pharmacoepidemiological Studies of Drug-Cancer Associations. Basic Clin Pharmacol Toxicol. 2018;122(5):451–459.

12. Pottegård A, Hallas J. New use of prescription drugs prior to a cancer diagnosis. Pharmacoepidemiol Drug Saf. 2017;26(2):223–227.

13. Huhn M, Nikolakopoulou A, Schneider-Thoma J, et al. Comparative efficacy and tolerability of 32 oral antipsychotics for the acute treatment of adults with multi-episode schizophrenia: a systematic review and network meta-analysis. The Lancet. 2019;394(10202):939–951.

14. Kinon BJ, Gilmore JA, Liu H, Halbreich UM. Prevalence of hyperprolactinemia in schizophrenic patients treated with conventional antipsychotic medications or risperidone. Psychoneuroendocrinology. 2003;28 Suppl 2:55–68.

15. Mortensen LQ, Andresen K, Burcharth J, Pommergaard H-C, Rosenberg J. Matching Cases and Controls Using SAS® Software. Frontiers in Big Data. 2019;Volume 2 - 2019.

16. Raghuthaman G, Venkateswaran R, Krishnadas R. Adjunctive aripiprazole in risperidone-induced hyperprolactinaemia: double-blind, randomised, placebo-controlled trial. BJPsych Open. 2015;1(2):172–177.

17. Stelmach A, Guzek K, Rożnowska A, Najbar I, Sadakierska-Chudy A. Antipsychotic drug—aripiprazole against schizophrenia, its therapeutic and metabolic effects associated with gene polymorphisms. Pharmacological Reports. 2023;75(1):19–31.

18. Sugai T, Suzuki Y, Yamazaki M, et al. Lower Prolactin Levels in Patients Treated With Aripiprazole Regardless of Antipsychotic Monopharmacy or Polypharmacy. J Clin Psychopharmacol. 2020;40(1):14–17.

19. Lillard JW, Jr., Moses KA, Mahal BA, George DJ. Racial disparities in Black men with prostate cancer: A literature review. Cancer. 2022;128(21):3787–3795.

20. Friedman Gary D, Habel Laurel A, Achacoso N, et al. Haloperidol and Prostate Cancer Prevention: More Epidemiologic Research Needed. The Permanente Journal. 2020;24(1):18.313.

21. Thomas JD, Longen CG, Oyer HM, et al. Sigma1 Targeting to Suppress Aberrant Androgen Receptor Signaling in Prostate Cancer. Cancer Res. 2017;77(9):2439–2452.

22. Redman B, Kitchen C, Johnson KW, Bezwada P, Kelly DL. Levels of prolactin and testosterone and associated sexual dysfunction and breast abnormalities in men with schizophrenia treated with antipsychotic medications. Journal of Psychiatric Research. 2021;143:50–53.

23. Ku SY, Gleave ME, Beltran H. Towards precision oncology in advanced prostate cancer. Nat Rev Urol. 2019;16(11):645–654.

24. Shigehara K, Izumi K, Kadono Y, Mizokami A. Testosterone and Bone Health in Men: A Narrative Review. J Clin Med. 2021;10(3).

25. Zarotsky V, Huang MY, Carman W, et al. Systematic literature review of the risk factors, comorbidities, and consequences of hypogonadism in men. Andrology. 2014;2(6):819–834.

26. Calabrese D, Giatti S, Romano S, et al. Diabetic neuropathic pain: a role for testosterone metabolites. J Endocrinol. 2014;221(1):1–13.

27. Allam JP, Bunzek C, Schnell L, et al. Low serum testosterone levels in male psoriasis patients correlate with disease severity. Eur J Dermatol. 2019;29(4):375–382.

28. Mangolim AS, Brito LAR, Nunes-Nogueira VDS. Effectiveness of testosterone replacement in men with obesity: a systematic review and meta-analysis. Eur J Endocrinol. 2021;186(1):123–135.

29. Yeap BB, Flicker L. Testosterone, cognitive decline and dementia in ageing men. Rev Endocr Metab Disord. 2022;23(6):1243–1257.

